# Retrospective Pooled Screening for SARS-CoV-2 RNA in late 2019

**DOI:** 10.1101/2020.05.14.20102079

**Authors:** Catherine A. Hogan, Natasha Garamani, Malaya K. Sahoo, ChunHong Huang, James Zehnder, Benjamin A. Pinsky

## Abstract

Reports have emerged documenting earlier SARS-CoV-2 cases than previously recognized. To investigate this possibility in the Bay Area, we retrospectively tested 1,700 samples from symptomatic individuals for the last 2 months of 2019. No SARS-CoV-2 positive pools were identified, consistent with limited transmission in this population at this time.

## Introduction

The emergence of severe acute respiratory syndrome coronavirus-2 (SARS-CoV-2) and the subsequent coronavirus disease 2019 (COVID-19) pandemic has revealed the critical importance of sentinel surveillance mechanisms for the rapid identification and response to new human pathogens. Phylogenetic analyses of SARS-CoV-2 suggest the virus first emerged weeks if not months before the World Health Organization (WHO) was notified of the original cluster of pneumonia cases associated with individuals who visited a wet market in Wuhan City, Hubei Province, in the People’s Republic of China (*1). Consistent with these estimates, an unconfirmed report in the lay press indicated the diagnosis of a COVID-19 case in Hubei Province on 17 November 2019 (*2*). Furthermore, retrospective testing in France detected SARS-CoV-2 in a respiratory sample collected December 27 2019, a month earlier than official cases were reported in the country (*3*). The San Francisco Bay Area presented among the first documented cases of community transmission in the U.S. (*4*), and a 6 February 2020 autopsy report from the Santa Clara County coroner confirmed detection of SARS-CoV-2 (*5*). Taken together, these reports suggest the possibility of earlier viral spread than previously recognized in Northern California. We recently employed a pooling strategy to leverage high-volume screening for community circulation of SARS-CoV-2 in individuals negative by routine respiratory virus testing during the first two months of 2020 (*4**). In this study, we extend pooled SARS-CoV-2 screening to include the last two months of 2019 to investigate possible early circulation of SARS-CoV-2 in this area.

## Methods

We performed a retrospective study that evaluated all available nasopharyngeal swab samples collected between 31 October 2019 to 31 December 2019 at Stanford Health Care from inpatients and outpatients that were negative by routine respiratory virus testing (Respiratory Pathogen Panel, Genmark Diagnostics or Xpert Xpress Flu/RSV, Cepheid) and that had not been tested for SARS-CoV-2. Pools were created by combining ten individual nasopharyngeal samples, and screening was performed using reverse transcriptase polymerase chain reaction (RT-PCR) targeting the nucleocapsid (*N2*) gene (*6*). This study was approved by the Stanford institutional review board (IRB), and individual patient consent was waived.

## Results

A total of 1700 individual nasopharyngeal specimens, corresponding to 170 pools, were included for SARS-CoV-2 testing. Of these, 841 samples were previously tested and negative by the Respiratory Pathogen Panel, and 859 were negative by the Xpert Xpress Flu/RSV. No SARS-CoV-2 positive pools were identified. The study time period corresponded to the onset of the 2019–2020 respiratory virus season with an increasing number of cases of influenza A, influenza B, and respiratory syncytial virus (RSV), and varying frequency of other seasonal viruses (Figure 1A 1B).

**Figure 1.**
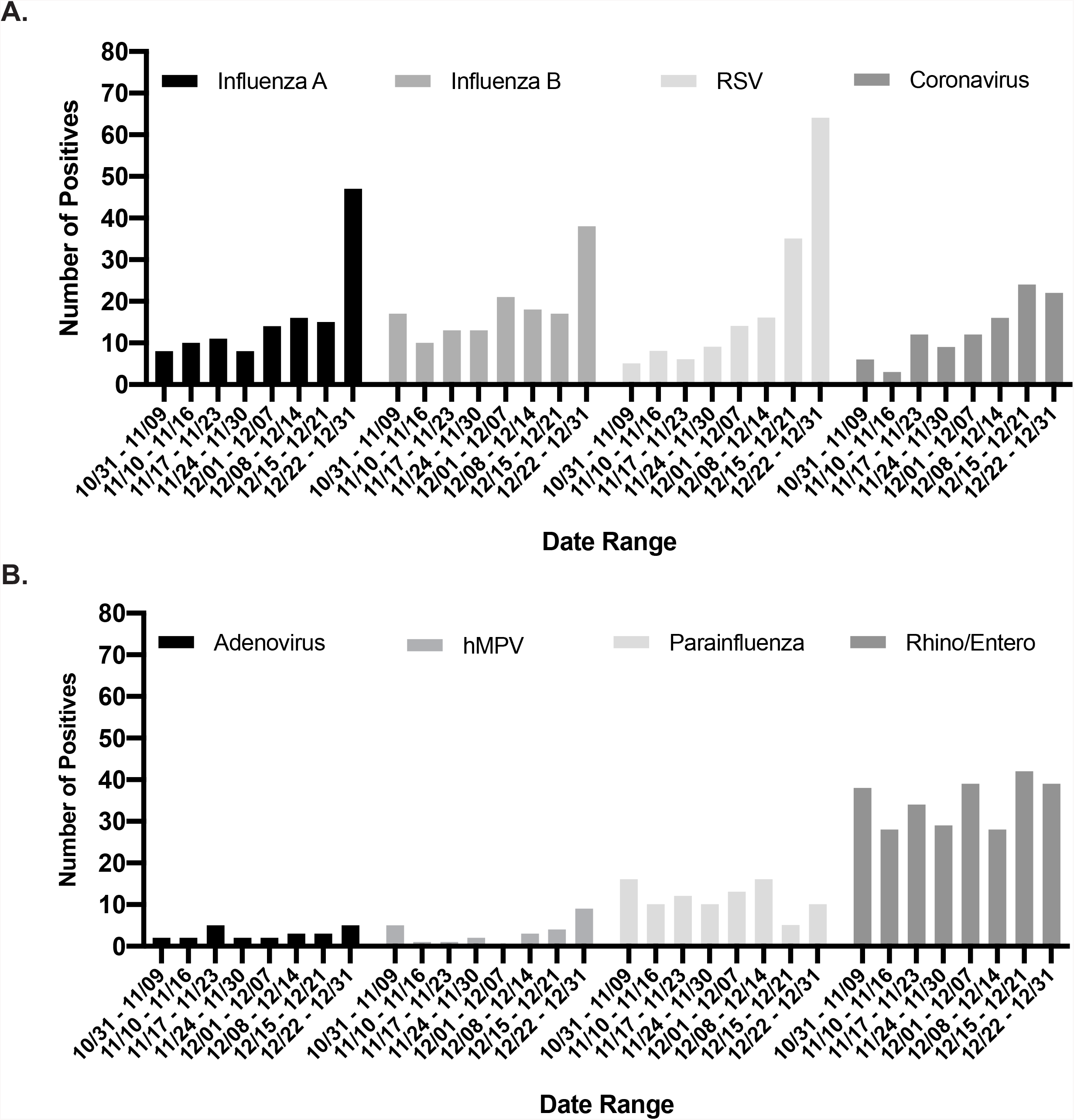
Respiratory surveillance November and December 2019. A) Influenza A, influenza B, respiratory syncytial virus (RSV), and seasonal coronaviruses, including HKU1, OC43, NL63, 229E. B) Adenovirus, human metapneumovirus (hMPV), parainfluenza viruses 1, 2, 3, and 4, and rhinovirus/enterovirus.

## Discussion

In this study, 1,700 nasopharyngeal samples collected from symptomatic individuals in the last two months of 2019 were screened by pooled testing, and no case of COVID-19 was detected. Phylogenetic analyses have estimated the emergence of SARS-CoV-2 to have occurred between 6 October 2019 and 11 December 2019 (*7*). Given this timeline, there has been growing interest to retrospectively identify “patient zero” in different geographic areas to better understand the spread of SARS-CoV-2, and to inform current and future surveillance strategies for emerging infectious diseases. Indeed, given the high volume of international travel prior to implementation of travel restrictions, travel-associated COVID-19 cases may have occurred in the U.S. earlier than previously recognized (*8*). However, monitoring for early community transmission of COVID-19 in the U.S was challenging due its similar clinical presentation to other respiratory virus infections, and overlap with the annual respiratory virus season. Strict indications for testing based on specific risk factors, coupled with limited testing capacity, further limited local COVID-19 case finding and contact tracing efforts (*9, 10*). Despite these shortcomings, this and previous work indicate that in the catchment area of our institution, symptomatic individuals without risk factors and diagnosed with SARS-CoV-2 infection began presenting to medical attention at the end of February 2020 (*4, 11*).

This study is limited by its sampling from a single institution, corresponding to a population that may not be representative of the underlying area as a whole. Further retrospective SARS-CoV-2 RT-PCR screening using specimens collected at other institutions throughout the U.S. will be necessary to fully understand early community transmission in this country. This work will complement phylogenetic viral sequence analysis and large-scale seroprevalence studies to characterize the regional and national emergence of SARS-CoV-2. Furthermore, it is possible that the use of pooled testing have led to lower sensitivity; however, pool sizes of 10 samples have been shown to maintain high performance compared to individual samples (*12*).

In summary, we employed a pooled screening strategy to investigate local community transmission of SARS-CoV-2 in late 2019 during the onset of the respiratory virus season. No COVID-19 cases were identified, consistent with limited transmission in this population at this time.

Dr. Hogan is an infectious diseases physician and medical microbiologist, currently a Visiting Instructor and a Global Health Diagnostics Fellow at the Stanford Department of Pathology. Her research interests include novel and point-of-care diagnostic methods, clinical impact of diagnostic methods and tropical medicine.

## Data Availability

The data generated and analyzed during the current study are available from the corresponding author on reasonable request.

